# Psychiatric side effects induced by chloroquine and hydroxychloroquine: a systematic review of case reports and population studies

**DOI:** 10.1101/2020.10.05.20207423

**Authors:** Fernanda Talarico, Sucheta Chakravarty, Yang S. Liu, Andrew Greenshaw, Ives Cavalcante Passos, Bo Cao

## Abstract

Chloroquine and hydroxychloroquine are commonly used drugs in the treatment of malaria as well as chronic diseases, such as rheumatoid arthritis, and systemic lupus erythematosus. Although various reports on possible psychiatric side effects of these drugs exist, the nature and extent of these effects remain poorly understood. Moreover, the relevance of these drugs in the treatment of early stages of COVID-19 necessitates a careful estimation of their side effects. Here, we provide a systematic review of the psychiatric side effects associated with chloroquine and hydroxychloroquine. We used PubMed, Scopus, and Web of Science platforms to identify relevant literature published between 1962 and 2020. Search terms included chloroquine, hydroxychloroquine, psychiatry, psychosis, depression, anxiety, bipolar disorder, delirium, and psychotic disorders. Only case reports and clinical trials were included. All studies included records of psychiatric side effects induced by either chloroquine or hydroxychloroquine or both. Both retrospective and prospective, randomized as well as non-randomized population studies were included. Overall, the psychiatric side effects are dose- and sex-independent. The most common psychiatric side effects reported are increased speech output/ excessive talking, increased psychomotor activity, irritable mood, auditory hallucinations, delusion of grandiosity, and suicide attempts, likely due to brain intoxicationbe of chloroquine or hydroxychloroquine. The symptoms can develop in a few hours to 11 weeks after drug intake and are normally reversed within a week after the drug withdrawal. We conclude that CQ and HCQ have the potential to induce psychiatric side effects. This study calls for further investigation of psychiatric symptoms induced by these drugs in the short and long term.

## Introduction

The World Health Organization (WHO) declared COVID-19 as a Public Health Emergency of International Concern on January 30th, 2020, and characterized it as a pandemic on March 11th, 2020, after its spread had reached several countries in Asia, Europe, and American continents. Shortly after, chloroquine (CQ) and hydroxychloroquine (HCQ) became two highly studied drugs for the treatment of early stages of COVID-19 (Chen Z. et al., 2020 - preprint; Tang W. et al., 2020). At least 20 clinical trials are being conducted (clinicaltrials.gov; search criteria: ‘condition’ - COVID-19 coronavirus SARS-CoV-2; ‘other terms’ - chloroquine hydroxychloroquine). While some studies have found positive results (Arshad S. et al., 2020; Chen Z. et al., 2020 - preprint), others report no such effect (Boulware D.R. et al., 2020; Mahevas M. et al., 2020 - preprint; Molina J.M. et al., 2020; Rosenberg E.S. et al., 2020). Additionally, adverse side effects such as rash development (Chen Z. et al., 2020 - preprint), headache (Chen Z. et al., 2020 - preprint), and cardiologic events (Mahevas M. et al., 2020 - preprint) were reported. Also, in the past, numerous studies have reported that CQ and HCQ can be associated with specific psychiatric side effects. However, these effects are not well understood.

CQ and HCQ have primary function as anti-inflammatory and immunosuppressive drugs, which, for the last two decades, have been primarily used as malaria prophylactic as well as for the treatment of malaria. They are also prescribed in rheumatoid arthritis (RA) and systemic lupus erythematosus (SLE) (Collins G.B. & McAllister M.S., 2008; Good M.I. & Shader R.I., 1982). The action mechanism includes inhibition of the acetylcholinesterase, inhibition of the prostaglandin synthesis, and prostaglandin E antagonism (Akhtar S. & Mukherjee S., 1993; Alisky J.M. et al., 2006; Bhatia M.S., 2012; Biswas P.S. et al., 2014; Hsu W.Y. et al., 2011; Mascolo A. et al., 2018). HCQ accumulates in the endosome, lysosomes, and Golgi vesicles leading to alkalinization of its content, which results in enzyme dysfunction. Then, the vesicles with viral content cannot enter the cell (Bhatia M.S., 1991; Bhatia M.S. et al., 1988; Bomb B.S. et al., 1975; Kabir S.M., 1969).

Although CQ and HCQ are considered safe drugs, they have a low safety therapeutic concentration margin (doses in which the drug achieve its efficacy without developing side-effects or toxicity), a long plasma half-life and terminal half-life (time required to reduce the drug concentration by one-half) at around one month (Aneja J. et al., 2019; Tango R.C., 2003). Therefore, adverse effects induced by these drugs can appear even within the therapeutic range and weeks after its administration (Aneja J. et al., 2019; Good M.I. & Shader R.I., 1982). The most well-known induced side effects are gastrointestinal problems, cardiac impairments, and cutaneous manifestation (Good M.I. & Shader R.I., 1982; Mascolo A. et al., 2018; Telgt D.S. et al., 2005). In addition, these medications can cross the blood-brain barrier reaching 10 to 20 times more concentration in the brain than in periphery blood plasma (Mascolo A. et al., 2018), leading to various psychiatric side effects such as behavioral symptoms, acute psychosis, depression, bipolar-like symptoms, anxiety, and suicide attempts (Akhtar S. & Mukherjee S., 1993; Biswas P.S. et al., 2014).

On April 4th, 2020, the US Food and Drugs Administration (FDA) issued an emergency use of HCQ to treat adults and adolescents hospitalized with COVID-19 and cannot participate in or do not have clinical trials available (https://www.fda.gov/media/136537/download). Although some psychiatric side effects were discussed in the document (“Psychosis, delirium, agitation, confusion, suicidal behavior, and hallucinations”), the contraindications mentioned were only for patients with retinal or visual field changes, and cardiac problems. However, previous studies looking at psychiatric side effects induced by CQ and HCQ suggest that these drugs should be prescribed with caution to patients with a previous diagnosis of mental illnesses or individuals with a family history of psychiatric disorders (Gonzalez-Nieto J.A. & Costa-Juan E., 2015; Good M.I. & Shader R.I., 1977; Lovestone S., 1991; Mascolo A. et al., 2018). Even though the FDA revoked its emergency use authorization on June 15th, 2020, it was evident that there is little information for physicians about the psychiatric side effects induced by HCQ and CQ. As there are still clinical-trials on these drugs being conducted, it is fundamental to bring up the attention for the induced psychiatric side effects. Therefore, the objective of this study is to focus on increasing the awareness of the CQ/HCQ induced psychiatric side effects by performing a systematic review of the topic. This will provide guidance of CQ/HCQ usage potentially not only on COVID-19, but other disorders.

## Methods

### Information sources, search strategy, and study selection

Studies were identified by searching four electronic databases in the following order: PubMed, Scopus, Web of Science, and Embase. The following Medical Subject Heading (MeSH) term filters were applied: (“Chloroquine” OR “Hydroxychloroquine”) AND (“Psychiatry” OR “Psychosis” OR “Depression” OR “Anxiety” OR “Bipolar Disorder” OR “Delirium” OR “Psychotic Disorders”). All four database searches were done in April 2020. A study was deemed relevant if it included a report of psychiatric side effects as well as a report of CQ or HCQ use. This encompassed both case reports and population studies. The relevance of a study was independently assessed by two authors (FT and SC) and the article contents were saved in an Excel spreadsheet. In case of a lack of agreement between these two authors, referral to a third author was planned. We excluded duplicate articles, articles that did not report any psychiatric side effects, did not report use of CQ or HCQ, were in vitro or animal studies, the full article was not available online, or was a comment to the journal (i.e., not an article). No language-based criterion was used for exclusion. We also excluded the review studies, since all of the relevant references in these reviews were either case reports or population studies that were already included (Alisky J.M. et al., 2006; Andrews R., 1985; Baird J.K., 2005; Berman J., 2004; Bhatia M.S. & Malik S.C., 1994; Bogaczewicz A. & Sobow T., 2017; Breuer O. & Schultz A., 2018; Chattopadhyay R. et al., 2007; Cooper R.G., 2008; Evans R.L. et al., 1984; Farhangian M.E. et al., 2015; Good M.I. & Shader R.I., 1977; Haładyj E. et al., 2018; Higgins G.C., 2018; Hu C. et al., 2017; Kalia S. & Dutz J.P., 2007; Khalifa A.E., 2007; Lewis J. et al., 2020; Mascolo A. et al., 2018; Mehat P. et al., 2017; Parker C., 2012; Peet M. & Peters S., 1995; Sato K. et al., 2020; Schlagenhauf P. & Steffen R., 2006; Steinhardt L.C. et al., 2011; Tango R.C., 2003; Taylor W.R.J. & White N.J., 2004; Wittes R., 1987).

### Data items and summary measures

For each included study we attempted to gather (1) demographic information about the patients (including age, gender and history of mental illness), (2) information on the use of CQ or HCQ (including the reason for drug use, if the drug was used alone or in combination with other drugs, its dose and duration), (3) characteristics of the psychiatric side effects (including the nature of the symptoms, whether it was self reported, onset time, treatment and time of remission). For population studies, the demographic information also included sample size, nationality and exclusion criteria for participants; the characteristics of psychiatric side effects included frequency and severity of the symptoms as well as an estimate of prevalence of psychiatric side effects, either from reports of percent of population affected or incidence rate ratios.

Apart from reporting the studies results, we also performed logistic and linear regressions to investigate which sex has greater chance of develping psychiatric side effects, and to observe whether there is a correlation between drug dosage and the psychiatric side effects, respectively. The analysis was done in R.

### Risk of bias across studies

The case-report and population studies differ on male/female rate, age (mean and range), medication dose, and the patient’s diagnosis (malaria, LSE, and RA). However, these disparities were not considered as a limitation since studies have reported that sex (Mohan D. et al., 1981) and age (Good M.I. & Shader R.I., 1977) are not predictors of psychiatric side effects induced by CQ or HCQ; and also the psychiatric side effects are not dose-dependent if prescribed within the therapeutic range (CQ and HCQ therapeutic dose: 6.5 mg/Kg/day) (Biswas P.S. et al., 2014; Collins G.B. & McAllister M.S., 2008; Mohan D. et al., 1981; Rollof J. & Vinge E., 1993).

## Results

### Study selection and characteristics

We identified 2,852 articles following the filters described above (294 articles on PubMed, 1,409 on Scopus, 241 on Web of Science, and 908 on Embase). After removing duplicate entries and applying our exclusion criteria, 57 eligible studies were included in the review (39 articles from PubMed, 16 articles from Scopus, none from Web of Science, and two from Embase) (Figure 1).

**Figure 1:**
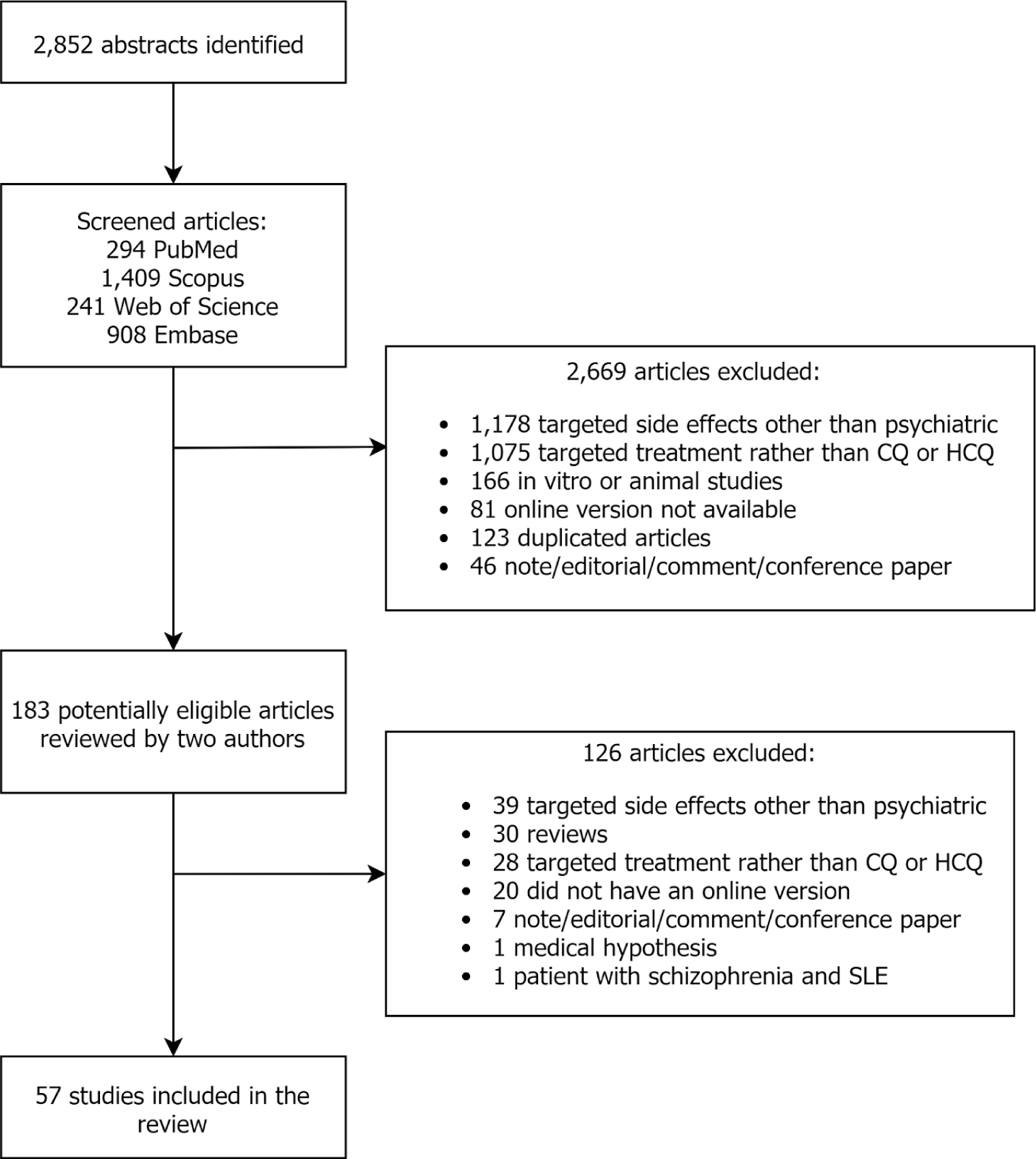
Flow diagram of the screening process

Among the 57 included studies, 38 were case reports and 20 were population studies. There was a lack of agreement on the content of nine articles between the two authors (FT and SC) highlighted by a third author (YL). A decision was made after discussion among the three authors. When mentioned, CQ and HCQ were prescribed for malaria, fever, systemic/discoid/cutaneous lupus erythematosus, rheumatoid arthritis, amebiasis, dermatitis, handicapping erosive plantar lichen, and as prophylactics after breast cancer with mastectomy.

### Studies overall

#### Clinical trials

Our search yielded a total of 20 population studies, of which, a study by Gacouin A. et al., (2017) was excluded as it only related to implications of deliberate self-poisoning by CQ. Table 1 lists the included population studies (N=19), alongwith the corresponding drug (CQ/HCQ) of relevance and properties specific to the drug group such as, sample size, dose, age and gender. This shows that CQ has been studied more frequently than HCQ and the drug group sample frequently included more women than men.

**Table 1.** Summary of population studies.

| Study | Drug | Sample size for each drug | Dose | Sex % (M / F) | Age |
| --- | --- | --- | --- | --- | --- |
| Frisk-Holmberg M. et al., (1979) | CQ | 100 | 0.25gm | more females overall | 55 (mean) |
| Boudreau et al., (1993) | CQ | 156 | 250mg | 100 / 0 | > 18 |
| Bhatia & Malik (1994) | CQ | 60 | 1 to 2.4gm | 30 / 70 | 6 to 36 |
| Barrett et al., (1996) | CQ+ | 1,181 | Not specified | 41.3 / 58.7 | Not specified |
| Petersen et al., (2000) | CQ - CQ+ | 1,223 - 1,827 | Not specified | Not specified | Not specified |
| Albright et al., (2002) | CQ | 22 | Not specified | 47 / 53 | ≤ 13 |
| Schlagenhauf et al., (2003) | CQ+ | 153 | 100mg CQ - 200mg proguanil | Not specified | Not specified |
| Meier et al., (2004) | CQ - CQ+ | 3,585 - 11,327 | Not specified | more females overall | Not specified |
| Korhonen et al., (2007) | CQ | 467 | Not specified | Not specified | Not specified |
| Schlagenhauf P. et al., (2009) | CQ+ | 135 | 100mg CQ - 200mg proguanil | Not specified | Not specified |
| Ursing J. et al., (2011) | CQ | 87 | 50mg/kg | 55 / 45 | 48 (mean) |
| Ribeiro N.P.O. et al., (2013) | HCQ | 31 | Not specified | 3.2 / 96.8 | 61 (mean) |
| Schneider et al., (2013) | CQ - CQ+ | Not specified | Not specified | more females overall | Not specified |
| Biswas et al., (2014) | CQ | 51 | 150mg | 52.9 / 47.1 | 31 (mean) |
| Sagara I. et al., (2014) | CQ+ - CQ+<sup>1</sup> | 113 - 114 | 600mg | 57.5 / 42.5<br>57.9 / 42.1 | 29.4 / 30.2 (mean) |
| Iudici M. et al., (2018) | HCQ | 83 | 5.3mg/kg | 5 / 95 | 41 (mean) |
| Mittal L. et al., (2018) | CQ - HCQ | 59 - 492 | 250mg CQ - 200 to 400mg HCQ | more females overall | 50 (mean) |
| Schneider A. et al., (2018) | CQ - CQ+ | 384 - 477 | Not specified | more males overall | Not specified |

CQ = Chloroquine; HCQ = Hydroxychloroquine; CQ+ = Chloroquine plus mefloquine or proguanil; +<sup>1</sup>: Chloroquine plus azithromycin; M = male; F = female; Sex is reported in percentage; Age is reported in years.

Multiple surveys were conducted on travellers who used these drugs as malaria prophylaxis (Albright T.A. et al., 2006; Barrett P.J. et al., 1996; Korhonen C. et al., 2007; Petersen E. et al., 2000). Typical components of these telephone, mail or web based surveys were name of the malarial prophylactic, compliance to the drug, nature and frequency of the side effects (see Table 2). These studies showed that on average, about 29% of the travellers using CQ (alone or with another drug) as prophylactic could experience some psychiatric side effects such as anxiety, depression, dizziness etc. However, whenever specified, the majority of these symptoms were reported to be mild in nature.

**Table 2:** Psychiatric side effects reported by the population studies with retrospective analysis.

| Study | Overall percentage* | Frequency (CQ ; CQ+) | Severity of symptoms |
| --- | --- | --- | --- |
| Barrett et al., (1996) # | 16% | Anxiety, depression, difficulty focussing, dizziness, hallucinations | 10.1% grade 1; 4.9 % grade 2; 0.9% grade 3; and 0.1% grade 4 |
| Petersen et al., (2000) | CQ: 15.5%;<br>CQ+ : 41.3% | Altered spatial perception (0.9%; 0.55%)<br>blurred vision (2.1%; 2.7%)<br>depression (1.3%; 2.6%)<br>dizziness (6.1%; 7.4%)<br>strange thoughts (0.5%; 1%) | CQ: 10% mild; 0.9% severe; 0.6% unacceptable effects; 2.8% changed medication.<br>CQ+: 32% mild; 4.6% severe; 1% unacceptable effects; over 5.9 changed medication |
| Albright et al., (2002) | 23% | Changes in sleep (14%)<br>headache (14%) | Not specified |
| Korhonen et al., (2007) # | 47% | *Anxiety, depression, dizziness, headache, insomnia, strange dreams, unsteadiness, visual disturbance and weakness | *49.5% mild; 40% moderate; 10.5% severe |

CQ = Chloroquine; CQ+ = Chloroquine plus proguanil; \*percentage of reported psychiatric side effects; #studies did not mention the frequency of the specific psychiatric side effects; severity grade 1 = relatively trivial; severity grade 2 = bad enough to interfere with daily activities; severity grade 3 = bad enough to make individuals seek medical advice; severity grade 4 = requiring hospital attention.

Schneider A. et al., (2018) also used a survey design but unlike the studies mentioned above, evaluated the relation between antimalarials (CQ, mefloquine, doxycycline etc.) use and overall physical and mental health of U.S. war veterans, using several independent questionnaires. 57.7% of the deployed (N=274) and 37.2% of the non-deployed (N=110) veterans, who reported taking CQ, were associated with mental health scores below the U.S. average. 18.9%, 12.8%, 10.4% and 11.4% of the deployed CQ users screened positive for PTSD, reported thoughts of death/self harm, other anxiety and major depression respectively. These proportions for the non-deployed CQ users were 7.4%, 9.2%, 2.5% and 4.5% respectively. However, these effects were better explained when deployment status and combat exposure were accounted for in the models.

Thus, despite the large sample size, results from the survey studies may need to be considered with caution, for some confounding variables may have been present. We revisit this in the discussion. However, more importantly, we also identified two nested case control and five randomized control studies, which offered better control over possible confounds. The nested case controls investigated risk of getting a first-time diagnosis of neuropsychiatric problems with the use of common antimalarials in a population of adults (more females) (Meier C.R. et al., 2004; Schneider C. et al., 2013). The results showed that the incidence rate for a first-time diagnosis of depression with intake of CQ (alone or with proguanil) could be as high as 7.6 per 1000 person-years. The same for a first time diagnosis of psychosis and panic attacks were 0.4 and 1.3 respectively (Meier C.R. et al., 2004). In Schneider C. et al. (2013), the overall incidence rate for diagnosis of anxiety, stress-related disorders, or psychosis with intake of CQ (alone or with proguanil) was 10.6 per 1000 person-years and the same for depression was 7.7. Also, women were at greater risk for developing these side effects than men. However, this was the only study to report such differences.

The randomized control trials also provide some evidence for psychiatric side effects induced by CQ (see Table 3). For example, Boudreau E. et al., (1993) found that overall (i.e.,when evaluations of side effects were combined across week 1 and weeks 9 to 12 of using CQ as prophylactic) 46% of the sample participants (N = 156, mean age = 25.7 years, all males) showed some CNS symptoms; insomnia and headache being most common (25% each), followed by dizziness (9%), irritability (5%), dreams (3%), anger (3%), moodiness (2%) and poor concentration (1%). Likewise, Schlagenhauf P., (2003) found 70% of the participants (N = 153) taking CQ (along with proguanil) to develop neuropsychological symptoms, such as, headache, strange or vivid dreams, dizziness, anxiety, depression, sleeplessness, and visual disturbance. However, 30% of such cases were moderate and only 6% were severe in nature. Later, an analysis of the mood profiles of the participants using the “Profile of Mood States” or POMS questionnaire showed that among CQ participants (N = 135), those below the median age (34 years) had more fatigue and tension than those at or above the median age. When used as a treatment rather than a prophylactic, Sagara I. et al., (2014) reported that CQ (along with azithromycin) induced psychiatric side effects like headache and dizziness in 13.2% and 9.6% of the sample population (N = 114) in a double-blind experiment. These proportions were even higher for the relevant sample in an open label experiment (N = 113, headache: 17.7%, dizziness: 15.9%, fatigue: 3.5%). On the other hand, Ursing J. et al., (2011) found no psychiatric side effects of CQ in treating malaria in children.

**Table 3:**
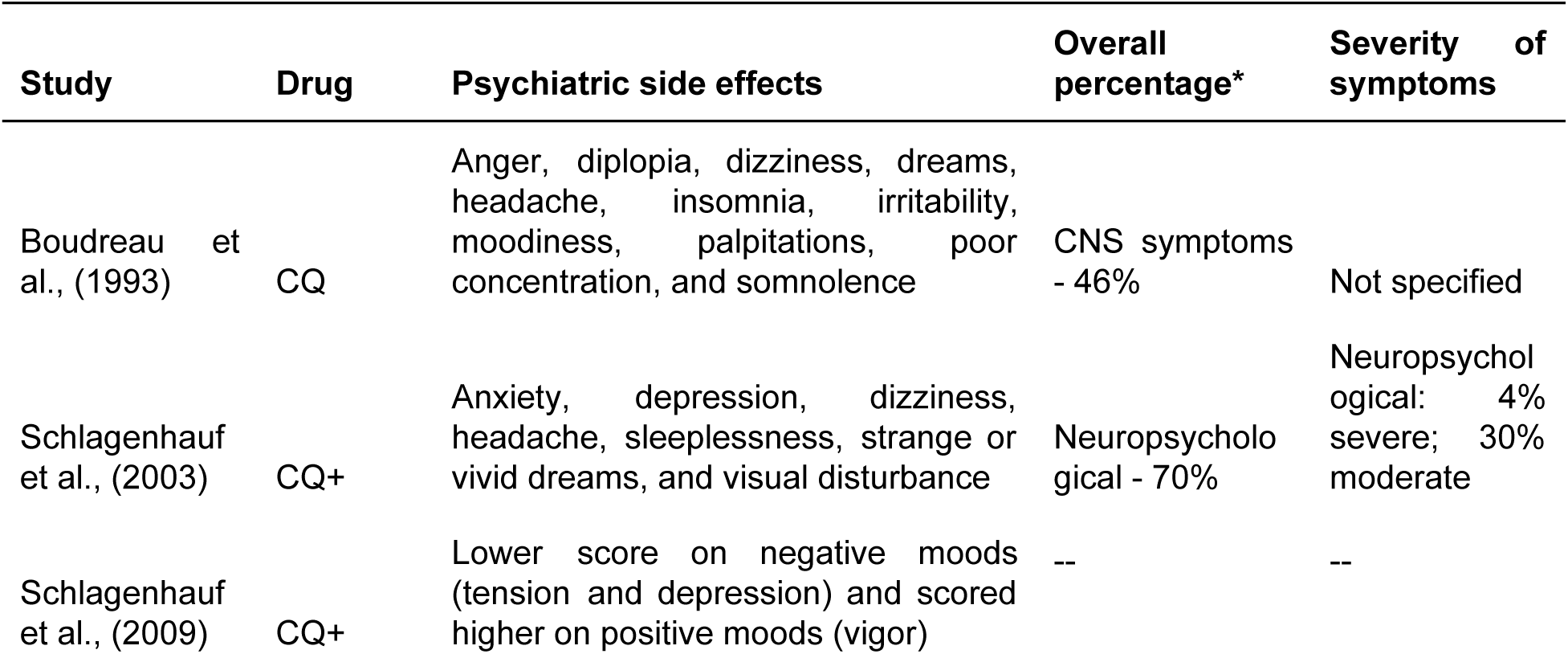
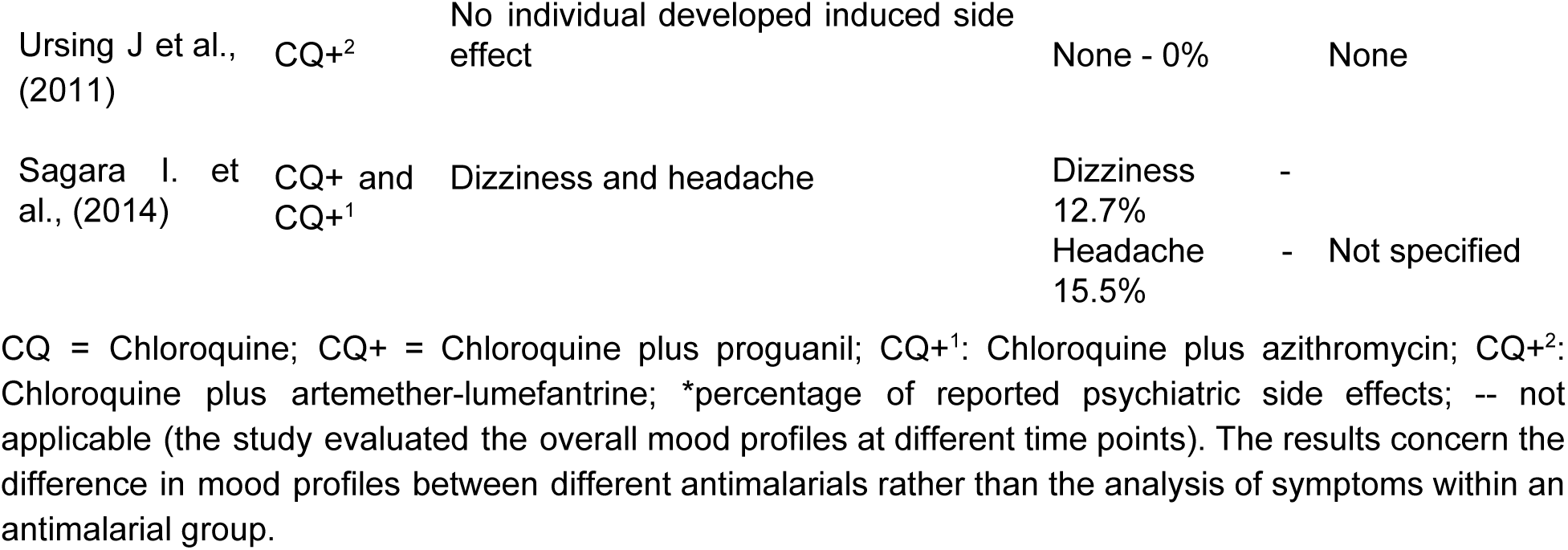
Psychiatric side effects reported by the randomized controlled trials.

Other studies suggest that within samples of patients with psychiatric side effects induced by CQ, organic psychosis was most common (53.2%), followed by schizophrenia-like symptoms (20%), mania (6.7%), depression (6.7%), anxiety (6.7%) and seizures (6.7%) (Bhatia M.S. & Malik S.C., 1994). Since psychiatric side effects induced by CQ seem to be short lived and in general disappear with CQ withdrawal, one study compared these effects of CQ to that of brief psychotic disorder (Biswas P.S. et al., 2014). In the psychosis following CQ patient group (N = 51, mean age = 31 years, 47% women) mixed affective psychosis was most common (76.2%), followed by depressive (14.28%), manic (4.76%), and non-affective (4.76%) psychosis. Also, prior marked stressors and abrupt onset of effects were less common in this group than the brief psychotic disorder group. Interestingly, psychosis following CQ patients showed more restlessness, agitation, irritability, anxiety, disturbed thought content, weight loss, visual hallucinations (with or without auditory hallucinations) and derealization. The psychosis following CQ group also showed less mannerism and posturing, motor retardation, blunted affect, and early insomnia and had better insight into their main and depressive symptoms. This study also reported no correlation between the amount of CQ consumed and the severity of the symptoms.

Finally, studies that looked into the side effects of CQ or HCQ as treatments for chronic diseases such as rheumatoid arthritis (RA), systemic lupus erythematosus (SLE), cutaneous lupus erythematosus (CLE) and dermatomyositis (DM), also report psychiatric side effects (see Table 4). For example, patients with RA who were treated with HCQ (N = 31, mean age = 61 years, 95% women) were at significant risk for anxiety, depression and suicidal ideation (Ribeiro N.P.O. et al., 2013). Small fractions of patients with CLE or DM (N = 532, mean age = 50 years, 85% women) who were treated with antimalarials (CQ, HCQ, quinacrine or a combination of these) developed psychiatric side effects such as dizziness (1.9%), headache (1.9%), sleep disturbances (0.9%) and psychosis (0.2%). The incidence rates for neurologic toxicities in general were 0.04 for HCQ and 0.02 for CQ (in person years) (Mittal L. et al., 2018). In patients with SLE being treated with HCQ (N=83, mean age = 41 years, 95% women), drug adherence was significantly less for those with better quality of life index. Non-adherent patients (29%) also reported less pain and higher general health. However, no significant difference existed between adherent and non-adherent patients in fatigue, anxiety, or depression (Iudici M. et al., 2018). In a sample of 100 patients (mean age = 55 years, more women), suffering from various chronic diseases such as RA, SLE etc., treatment with CQ produced side effects in 15% of the sample. Psychiatric side effects included visual disturbances, headache, apathy, anxiety, and fatigue (Frisk-Holmberg M. et al., 1979). Also, there was a positive correlation between the serum concentration of CQ and the side effects. However, there was no such trend between the dose of CQ and side effects.

**Table 4:**
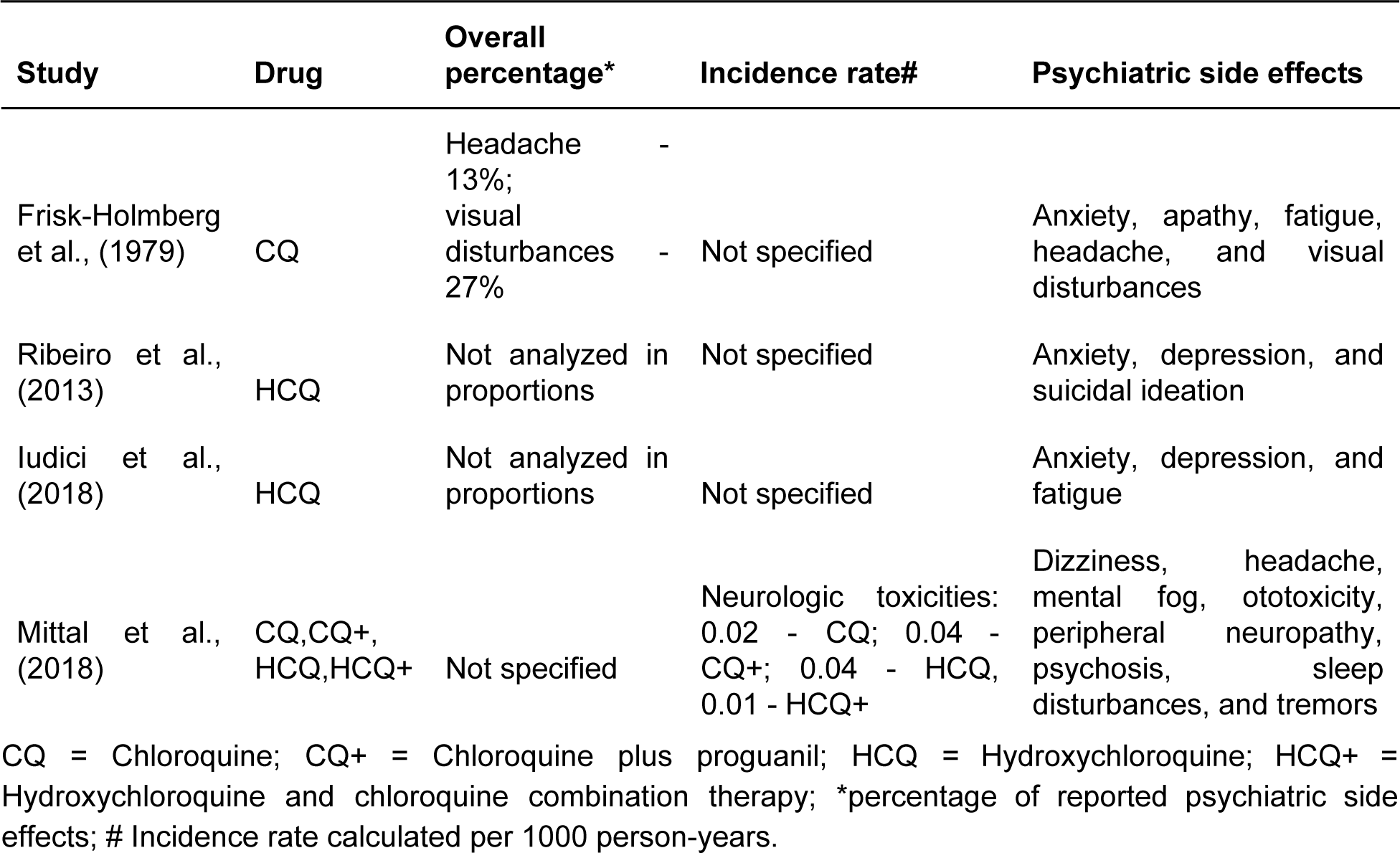
Psychiatric side effects induced by chloroquine or hydroxychloroquine for the treatment of chronic diseases.

#### Case - reports

There were 62 cases reported from the 38 studies. 56 cases were drug-induced psychiatric side effects and five cases were inconclusive: 1) individual relapsed into a manic episode without being further exposed to CQ (Akhtar S. & Mukherjee S., 1993 - 2nd case report); 2) individual did not develop psychiatric side effects after reintroduction of CQ (Akhtar S. & Mukherjee S., 1993 - 3rd case report); 3) individual was consuming alcohol and was a chronic CQ user (Davis T.M. & Ilett K.F., 2000); 4) authors reported an inconclusive diagnosis (Sharma S. et al., 2016); 5) individual was found dead in his room due to suicide by multiple knives stabs to the head (Jousset N. et al., 2010). Although a toxicological examination confirmed the recent intake of CQ and mefloquine, it is hard to draw a conclusion. One case was further excluded as it did not use CQ or HCQ as treatment (Sapp O.L., 1964 - 1st case report).

The first reported cases of psychosis and mania in adults was in the 1960s (Dornhorst A.C. & Robinson B.F., 1963; Mustakallio K.K. et al., 1962; Rab S.M., 1963), while the first reported cases in children was in 1988 (Halder D. et al., 1988). The majority of cases did not have a family history of any psychiatric disease. One individual had a diagnosis of borderline personality disorder when started the treatment (Gonzalez-Nieto J.A. & Costa-Juan E., 2015), and one has a mother and an uncle diagnosed with bipolar affective disorder (Akhtar S. & Mukherjee S., 1993). The summary of the case-reports is presented in Table 5.

**Table 5.** Summary of case-reports.

|  | Hydroxychloroquine | Chloroquine<br>(or CQ+*) | Total |
| --- | --- | --- | --- |
| Induced psychiatric side effect<br>(number of cases) | 9 | 53 | 62 |
| Reason for the prescription<br>(number of cases) |  |  |  |
| Malaria | 0 | 15 | 15 |
| Fever | 1 | 12 | 13 |
| SLE or DLE | 5 | 4 | 9 |
| RA | 3 | 3 | 6 |
| Amebiasis | 0 | 3 | 3 |
| Dermatitis | 0 | 1 | 1 |
| Lichen | 1 | 0 | 1 |
| Breast cancer | 0 | 1 | 1 |
| Dose (mean) | 275 mg | 877 mg | 750 mg<br>(100mg to 4.7g) |
| Sex (M / F) | 02 / 07 | 29 / 22 | 31 / 29 |
| Age in years<br>(mean, younger - older) | 54.1 (17 - 80) | 27.9 (1 - 72) | 32.6 (1 - 80) |

\*CQ+: chloroquine (CQ) plus mefloquine, proguanil, artesunate and/or NSAIDs; SLE = systemic lupus erythematosus; DLE = discoid lupus erythematosus

The mean of psychiatric side effect onset was 9.3 days (from 3 hours to 12 weeks) after drug intake (calculated excluding individuals taking HCQ for 8 and 6 years) (Hsu W.Y. et al., 2011; Kwak Y.T. et al., 2015). As previous studies reported that psychiatric side effects are dose-independent (Biswas P.S. et al., 2014; Collins G.B. & McAllister M.S., 2008; Mohan D. et al., 1981; Rollof J. & Vinge E., 1993), we performed a linear regression using the CQ and HCQ doses as the dependent variable, and the days until psychiatric side effects onset as the independent variable to observe whether there is a correlation between them. For this analysis we excluded an outlier (dosage was higher than 3 standard deviation) as we believed that this individual was influencing the results (Figure S1). We did not observe any effect of dose on the onset of psychiatric side effects in days using all individuals nor stratifying for sex (Figure 2).

**Figure 2:**
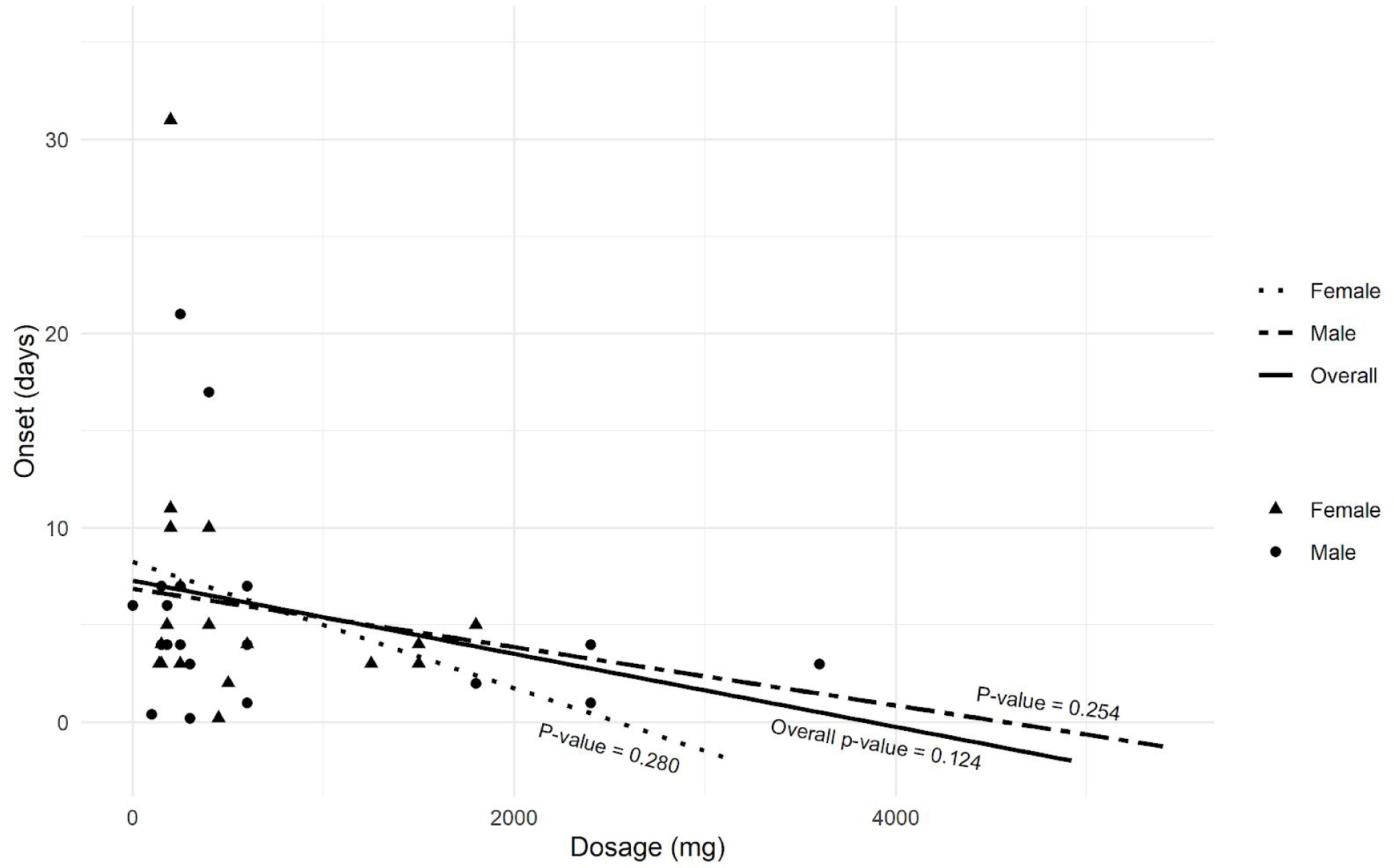
Distribution of drug dosage and the onset of psychiatric side effects with all case-reports excluding one report in which the patient took high drug dosage (4,700 mg). P-value: result of linear regression.

A total of 57 psychiatric side effects was mentioned (Table S1). The most common were excessive talking, increased psychomotor activity, auditory hallucination, and decreased need for sleep / insomnia. When performing a logistic regression using psychiatric side effects as the dependent variable and sex as the independent variable, we did not find statistical differences in most of the frequency of psychiatric side effects between females and males (p-value = 0.13), as previously reported (Mohan D. et al., 1981). However, women appear to have higher probability of developing suicidal ideation (p-value = 0.032) (Figure 3).

**Figure 3:**
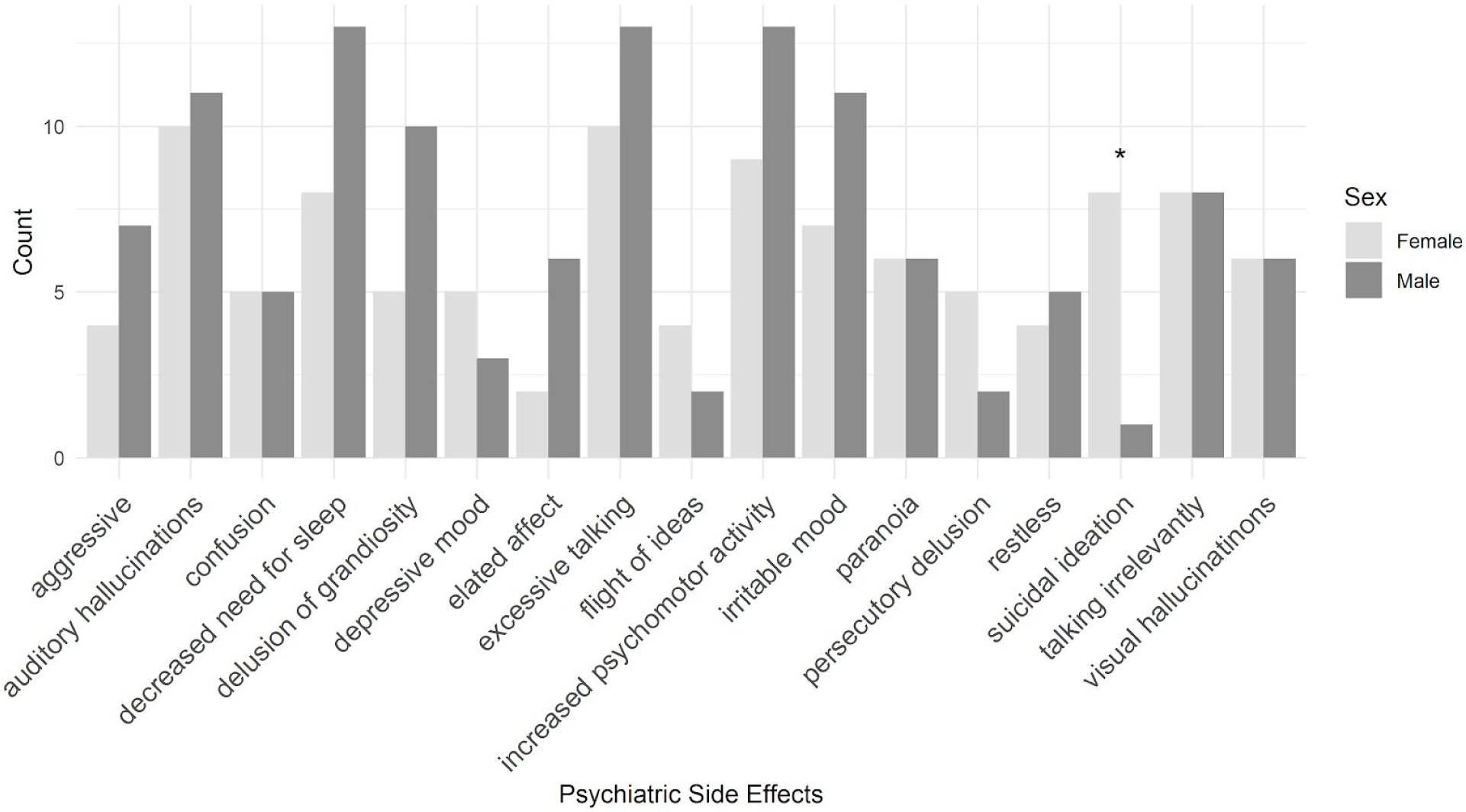
Frequency of the top psychiatric side effects by sex. * logistic regression p-value = 0.032.

The symptoms may develop once CQ or HCQ is reintroduced. Six individuals suffered from psychiatric side effects at different time-points: 3 years (Bhatia M.S., 2012), 15 months (Akhtar S. & Mukherjee S., 1993 - 3rd case report), and 5 years (Brookes D.B., 1966) before the case-report; two weeks (Manzo C. et al., 2017), three weeks (Plesnicar B. et al., 2013), three months (Kwak Y.T. et al., 2015), and 5 years (Gonzalez-Nieto J.A. & Costa-Juan E., 2015) after the first case. Differently, some individuals had taken CQ or HCQ before the case-report without having any complications (Cras P. & Martin J.J., 1990; Gulec G. et al., 2009; Rollof J. & Vinge E., 1993). In addition, two individuals experienced recurrences several months after taking the last dose of CQ (Regan E. et al., 1985).

Usually, the symptoms disappear without the need of treatment within seven days after the drug withdrawal due to its half-life, but it can persist for several weeks in case of overdosage (1,500mg in a 7 years-old girl) (Zaki S.A. et al., 2009). However, medications are often prescribed to relieve the symptoms (Ward W.Q. et al., 1985). Antipsychotics or antidepressants were the treatment of choice for psychiatric side effects in 47 cases and electroconvulsive therapy in two (Akhtar S. & Mukherjee S., 1993), while in 11 cases drug withdrawal was sufficient to ameliorate the symptoms. The most prescribed medications were tricyclic antidepressants, haloperidol, risperidone, and diazepam. The remission time varied between one day to five months.

## Discussion

We conducted a systematic review of the psychiatric side effects of CQ and HCQ. To our knowledge, this study is the very first of its kind. Notably, the majority of relevant studies were case reports and randomized control trials are very few in this topic. Thus, it is difficult to draw conclusions with greater confidence. Accordingly, we focus on trends in the results that are consistent across multiple studies. These trends not only help better understand the potential risks of these drugs but also offer plausible hypotheses to be tested in future with better control conditions.

Overall, the population level studies available on this literature suggest that there is a small to moderate probability of experiencing short term psychiatric side effects such as anxiety, depression, dizziness, headache, sleep disturbances etc., with the use of CQ/HCQ, either as a malaria prophylactic or as treatment for malaria infection or as treatment for other relevant chronic diseases such as, RA, SLE etc. These side effects are in general mild in nature. Notably, these studies frequently consisted of the adult population with more females. There is no clear evidence that onset of these symptoms is dose dependent. In comparison to other psychiatric episodes such as brief psychotic disorder, cases of psychiatric side effects induced by CQ may be more prominent in nature but at the same time, the patients also show better insight into these symptoms.

Although the frequent use of CQ as malaria prophylactic makes the large scale surveys of side effects done on travellers highly relevant and important, we speculate multiple confounds that warrants caution. For example, participation in the surveys was voluntary, thus the response rates varied and it is possible that users who experienced symptoms were more likely to respond. These studies also differed on the time (relative to travel) when the surveys were conducted and the participants were often prompted with a list of psychiatric side effects of the drug as previously reported in the literature. These factors could have also biased some of the responses. For example, travellers who did not take any antimalarial also reported some psychiatric side effects in Petersen et al. (2000). Also, in case of chronic diseases like RA, SLE etc., a careful consideration may be required to estimate if the side effects were produced by the drug or resulted from the disease itself.

Accordingly, we assign greater impact to the randomized control studies, which in most cases were double blind and thus less prone to confounding variables. The results from these studies were also in favour of the idea that with the exception of children, short term psychiatric side effects induced by CQ is probable for a moderate proportion of the sample and probably even more so when combined with another antimalarial such as proguanil. However, in line with our speculation that self reports of the side effects could also be due to reasons other than CQ intake, Schlagenhauf et al. (2003) found that even participants in the placebo group reported some of these symptoms. Nested case control design studies may also be more reliable than survey studies, as it provides superior unbiased estimates of the effects. Participants with specific symptoms were first identified and exposure to the drug was evaluated. Then, for each relevant case, multiple control patients were selected at random from the same database. These also showed evidence for short term psychiatric side effects induced by CQ/HCQ, albeit with small incidence rates.

Further, a careful estimation of the case reports, which included controlling for confounds such as previous history of mental illness, overdose etc., showed that CQ and HCQ may indeed give rise to short term psychiatric side effects. Most psychiatric side effects developed within 3 hours to 1 week after the drug intake. However, some patients had a long term effect, where six individuals developed the symptoms after a month and two individuals after taking the drug for 8 and 6 years respectively. Normally, the psychiatric side effects ceased without the need of treatment (i.e. with the withdrawal of the drug), but sometimes antipsychotic were prescribed to alleviate the symptoms. Multiple cases also showed recurrence of the symptoms with the reintroduction of the drugs. Together, these trends suggest that the psychiatric symptoms are more likely to be causal to the intake of CQ or HCQ than the underlying condition or reason for drug intake (for example fever, chronic disease etc.). A linear regression analysis between the onset time for psychiatric symptoms and the dose of the drug showed a negative trend, i.e., higher doses were associated with faster onset of symptoms. However, this trend failed to reach significance. When considering the frequency of the different psychiatric side effects, mania-like symptoms are more common, such as excessive talking, increased psychomotor activity, auditory hallucination, and decreased need for sleep or insomnia were more common. Gender-based differences in the frequency of symptoms were not significant, except for women reporting suicidal ideation significantly more often than men.

Taken together, there is considerable reason to believe that short term psychiatric side effects induced by CQ/HCQ is a possibility. The most discussed risk factors are family history, previous psychiatric disorders, and age, but the majority of cases reported happened in individuals without a history of psychiatric diseases. Elderly patients are at higher risk of developing such side effects due to changes in pharmacokinetics and pharmacodynamics. Nonetheless, due to the limitations brought up regarding the voluntary response in survey data, self-reports side effects, and placebo group reporting similar symptoms than case group, populational level studies addressing these limitations is needed as well as more investigation to understand the psychiatric side effects nature and extent.

Below we discuss the possible mechanisms for the occurrence of these symptoms with the intake of CQ/HCQ.

### Prevalence and risks of psychiatric side effects induced by CQ or HCQ

There are inconsistencies among studies on the prevalence of psychiatric side effects induced by CQ/HCQ. In 2015, Gonzalez-Nieto JA and Costa-Juan E calculated a prevalence of 0.09%, while Mascolo A and co-workers in 2018 reported 1% and 7% prevalence of neuropsychiatric disorders and psychiatric disorders, respectively. Although having a small sample size, other studies reported that 43% of patients treated with CQ for RA developed depression (Drew J.F., 1962; Mohan D. et al., 1981) and 29% of patients taking HCQ for RA developed mild to severe depression (Wilkey I.S., 1971). Some studies found a higher prevalence among women in menopausal age (Drew J.F., 1962), while in others the frequency was higher in children (Akhtar S. & Mukherjee S., 1993). Other two studies calculated the incidence rate (IR) (Table 6).

**Table 6:** Incident rate (IR) calculated by several psychiatric side effects in two studies.

| Study | Psychiatric side effect | Incident rate (IR)* |
| --- | --- | --- |
| **Schneider et al., 2013** | Anxiety or stress-related disorders or psychosis | 10.6 |
|  | Depression | 7.7 |
| **Meier et al., 2004** | Depression | 7.6 |
|  | Psychosis | 0.4 |
|  | Panic attack | 1.3 |

\*Incidence rate calculated per 1000 person-years.

Although studies suggested family history or presence of previous psychiatric disorders as the major predisposing factor (Collins G.B. & McAllister M.S., 2008; Gonzalez-Nieto J.A. & Costa-Juan E., 2015; Schneider C. et al., 2013), most of the CQ/HCQ-induced psychiatric side effects reported in this review occurred in people without a history of psychiatric diseases. However, it is important to mention that individuals with prodromal mental health symptoms, such as difficulty concentrating, anxiety, and nightmares, might have increased chances of developing psychiatric side effects (Ferraro V. et al., 2004). In addition, elderly patients also appear to have higher probability of such side effects. They present changes in pharmacokinetics and pharmacodynamics leading to an enhanced incidence of drug toxicity and, consequently, side effects. In addition, HCQ and CQ tend to accumulate more easily in the elderly’s fat tissues due to the diminished water in their body (Altintas E., 2015; Kwak Y.T. et al., 2015. Therefore, symptoms may be present for a longer period of time and even in lower doses than in adults and children (Kwak Y.T. et al., 2015).

Other possible risk factors are patients with auto-immune infectious disease (Gonzalez-Nieto J.A. & Costa-Juan E., 2015), epilepsy (Schneider C. et al., 2013), hepatic insufficiency (Sahoo S. et al., 2007), hypoalbuminemia (Gonzalez-Nieto J.A. & Costa-Juan E., 2015), smoke (Meier C.R. et al., 2004), alcohol intake (Gonzalez-Nieto J.A. & Costa-Juan E., 2015), low BMI (Gonzalez-Nieto J.A. & Costa-Juan E., 2015; Schneider C. et al., 2013), and co-administration of drugs that interfere with the CYP450 enzymes family (Gonzalez-Nieto J.A. & Costa-Juan E., 2015; Maxwell N.M. et al., 2015; Rollof J. & Vinge E., 1993).

The CYP450 enzyme family plays an important role in the metabolism of drugs in humans (Tornio A. & Backman J.T., 2018). CYP1A1, CYP1A2, CYP2C8, CYP2C1, CYP2D6, and CYP3A4 enzymes are believed to be involved in CQ and HCQ metabolism (Maxwell N.M. et al., 2015). Genetic variants, such as single nucleotide polymorphisms (SNPs), in genes that codify those enzymes, might be one of the causes of CQ or HCQ intoxication. For example, reduced CYP2D6 activity may cause serious and prolonged CQ brain intoxication within normal doses (Maxwell N.M. et al., 2015). Then, the co-administration of drugs that interact with CYP enzymes can reduce or increase its activity. One case-report showed that co-administration of CQ and fluoxetine, a CYP2D6 inhibitor, might have diminished CQ metabolism in the brain, eliminating the psychiatric side effects (Maxwell N.M. et al., 2015). In other cases, the administration of drugs that interact with CYP enzymes, such as naproxen, rifampin, and ciprofloxacin, might have contributed to the induced psychiatric side effects (Mohan D. et al., 1981; Rollof J. & Vinge E., 1993).

A better knowledge of CQ/HCQ pharmacological mechanisms is fundamental to improve the understanding of how they induce psychiatric side effects. Consequently, we would have better insights on the prevention and treatment of these conditions (Bogaczewicz J. et al., 2014).

### Mechanism of how the psychiatric conditions may develop

The CQ/HCQ induced-psychiatric side effects might be due to the drug’s ability to cross the blood-brain barrier. In the brain, they can have a concentration 10 to 20 times higher than in plasma (Mascolo A. et al., 2018). The complete drug absorption occurs within 2 to 4 hours after its oral administration. CQ plasma concentration can reach 53% of its peak concentration 5 days after the last dose, and detectable amounts can be found even 42 months after its withdrawal (Sapp O.L., 1964). The individuals’ differences in absorption and degradation, and the consequent differences in the drug steady-state concentration may lead to the variability in the time window between the drug intake and psychiatric side effect appearance (Altintas E., 2015; Garg P. et al., 1990; Manzo C. et al., 2017; Sapp O.L., 1964).

Several studies described an imbalance in the dopaminergic pathway after the drug intake. HCQ causes neurotoxicity (Bhatia M.S., 2012; Bogaczewicz A. et al., 2016; Hsu W.Y. et al., 2011; Sahoo S. et al., 2007), NMDA excitotoxicity, and GABA inhibition (Aneja J. et al., 2019; Hsu W.Y. et al., 2011; Sahoo S. et al., 2007). So, the dopamine activity is enhanced causing psychosis (Akhtar S. & Mukherjee S., 1993) and mania-like symptoms (Mohan D. et al., 1981). CQ and HCQ also cause a disturbance of the cholinergic system (Akhtar S. & Mukherjee S., 1993; Bhatia M.S., 2012; Biswas P.S. et al., 2014; Collins G.B. & McAllister M.S., 2008) related to the inhibition of acetylcholinesterase (Bogaczewicz A. et al., 2016; Mohan D. et al., 1981) and blockage of the muscarinic cholinergic receptors (Hsu W.Y. et al., 2011). The serotonin transporter is inhibited, elevating the levels of the neurotransmitter in the synapsis, causing mania and psychosis (Alisky J.M. et al., 2006).

In addition, there is a reduction of the prostaglandin E (Akhtar S. & Mukherjee S., 1993; Bhatia M.S., 2012; Biswas P.S. et al., 2014; Collins G.B. & McAllister M.S., 2008), and a down-regulation of glycoprotein-P in the blood-brain barrier (Alisky J.M. et al., 2006). This protein is responsible for removing a variety of substances from the neurons and clearance of antidepressants across the brain-blood barrier. Therefore, if a patient is taking CQ and antidepressants, it could lead to a mental status change. Finally, CQ increases the EEG frequencies acting as a cerebrocortical stimulant (Biswas P.S. et al., 2014), but decreases voltage, which may lead to seizures. We thereby hypothesize that CQ and HCQ besides inducing psychiatric side effects, may be risk factors for neurological disorders like seizure and Alzheimer’s Disease, especially as a long term effect.

### Limitations

Our study has some limitations. Since most of the published articles on this topic are case-reports, one may argue that it is difficult to draw a solid conclusion about the risk factors and the prevalence of psychiatric side effects. However, we reviewed a considerable number of cases and all of them controlled for confounding factors, such as previous history of mental illness and overdose. In addition, most of populational studies have a good sample size (more than 100 individuals), in which seven were randomized control studies or nested case control studies, that accounted for confounding variables and provide unbiased estimates of the effects (Albright T.A. et al., 2006; Barrett P.J. et al., 1996; Biswas P.S. et al., 2014; Boudreau E. et al., 1993; Gacouin A. et al., 2017; Korhonen C. et al., 2007; Meier C.R. et al., 2004; Mittal L. et al., 2018; Petersen E. et al., 2000; Ribeiro N.P.O. et al., 2013; Sagara I. et al., 2014; Schlagenhauf P., 2003; Schlagenhauf P. et al., 2009; Schneider A. et al., 2018; Schneider C. et al., 2013; Ursing J. et al., 2011). Therefore, we believe that we overcome the limitation mentioned above and our conclusion should be taken into consideration.

## Conclusion

The studies reviewed in this article suggest short term psychiatric side effects induced by CQ/HCQ. However, due to some inconsistencies regarding the risk factors to develop CQ/HCQ-induced psychiatric side effects and the limitations brought up above, more studies addressing these limitations is needed as well as more investigation to understand the psychiatric side effects nature and extent. Most importantly, clinicians should be extra careful with the potential development of psychiatric side effects in individuals being tested for CQ and HCQ for the treatment of COVID-19.

## Data Availability

All studies included in this review are cited in the manuscript. No additional data were used.

