## Supplemental Table and Figure 1 for "Psychiatric side effects induced by chloroquine and hydroxychloroquine: a systematic review of case reports and population studies"

**Figure S1**: Distribution of drug dosage and the onset of psychiatric side effects with all case-reports. Arrow: highlighting the individual who took high drug dosage (4,700mg). P-value: result of linear regression.

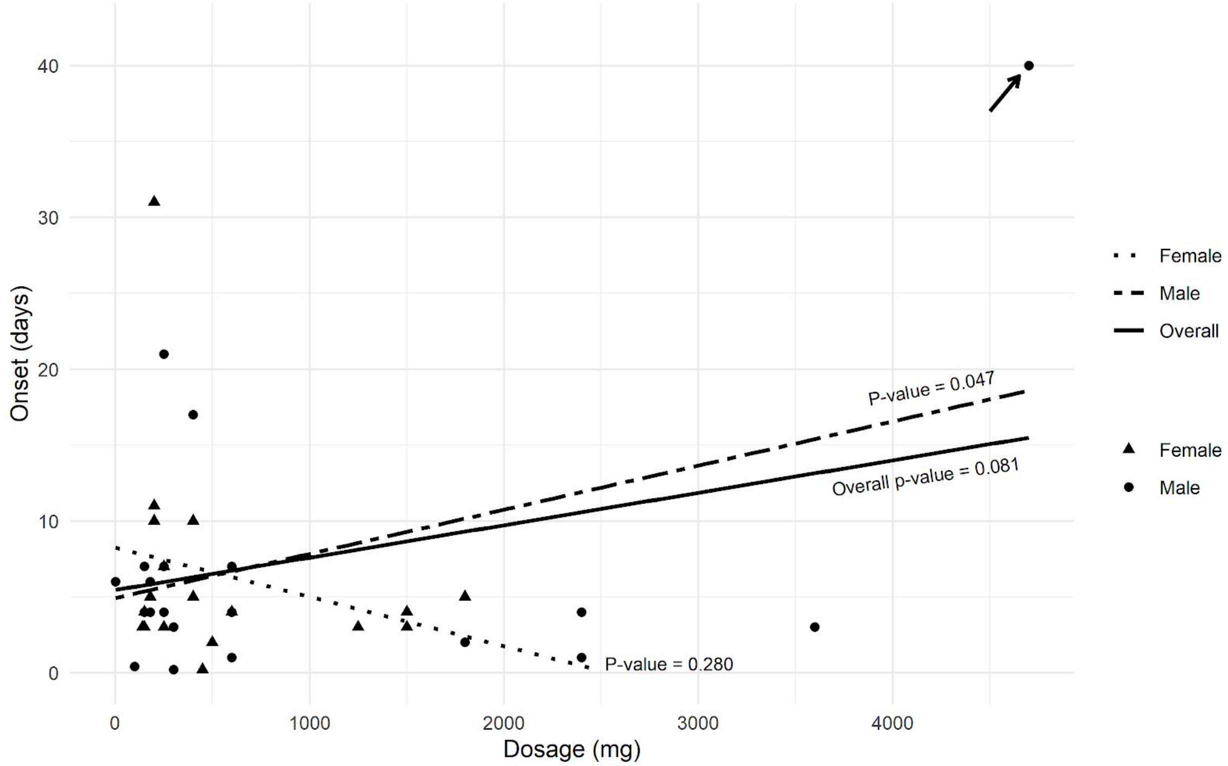

**Table S1:** Psychiatric side effects in case-reports studies.

| Psychiatric side effects | How many appearances |
|---|---|
| excessive talking | 23 |
| increased psychomotor activity | 22 |
| auditory hallucination | 21 |
| decreased need for sleep / insomnia | 21 |
| irritable mood | 18 |
| talking irrelevantly | 16 |
| delusion of grandiosity | 15 |
| paranoia | 12 |
| visual hallucination | 12 |
| aggressive/violence | 11 |
| confusion | 10 |
| restless | 9 |
| suicidal ideation | 9 |
| depressive mood | 8 |
| elated affect | 8 |
| persecutory delusion | 7 |
| flight of ideas | 6 |
| distractible | 5 |
| generalized anxiety | 5 |
| non-specific psychotic symptoms | 5 |
| non-specific delusion | 4 |
| catatonic state | 3 |
| derealization/depersonalization | 3 |

| | |
|---|---|
| disturbed appetite | 3 |
| headaches | 3 |
| laughing without reason | 3 |
| lightheadednes | 3 |
| tactile hallucination | 3 |
| tremor | 3 |
| worthlessness | 3 |
| abusive | 2 |
| delirium | 2 |
| increased energy | 2 |
| increased self-esteem | 2 |
| kinaesthetic hallucination | 2 |
| loss of interest | 2 |
| lost of memory | 2 |
| weeping spells | 2 |
| cheerful | 1 |
| crying spells | 1 |
| delusion of erotomania | 1 |
| delusion of reference | 1 |
| excitation | 1 |
| expansive mood | 1 |
| feel being lost | 1 |
| hostility | 1 |
| increased libido | 1 |
| non-specific hallucination | 1 |

| | |
|---|---|
| non-specific manic symptoms | 1 |
| over socializing | 1 |
| partition delusion | 1 |
| pessimism | 1 |
| psychomotor retardation | 1 |
| psychotic episode | 1 |
| racing thoughts | 1 |
| self-harm | 1 |
| sweating | 1 |
